# Pre-infusion plasma proteomics identifies an endothelial–immune priming signature predictive of severe cytokine release syndrome and neurotoxicity following CAR T-cell therapy in relapsed/refractory lymphoma

**DOI:** 10.64898/2026.03.29.26349664

**Authors:** Ehsan Irajizad, Johannes F. Fahrmann, Hiroyuki Katayama, Paolo Strati, Ranjit Nair, Dai Chihara, Sairah Ahmed, Swaminathan P. Iyer, Frederick L. Locke, Marco Davila, Christopher R. Flowers, Elizabeth Shpall, Robert Jenq, Sattva S. Neelapu, Samir Hanash, Jason Westin, Michael D. Jain, Teny M. John, Neeraj Y. Saini

## Abstract

**Background:** Severe cytokine release syndrome (CRS) and immune effector cell-associated neurotoxicity syndrome (ICANS) remain frequent, life-threatening complications of CD19 chimeric antigen receptor (CAR) T-cell therapy and constrain its safety, scalability, and outpatient adoption. Existing predictive models lack sufficient external validation for routine clinical use, and pre-infusion biomarkers that capture host susceptibility before infusion are urgently needed.

**Methods:** We applied unbiased mass-spectrometry-based proteomics to pre-infusion biofluids from 98 prospectively-followed adults with relapsed/refractory (r/r) lymphoma at two academic centers (MD Anderson Cancer Center, n = 39, plasma; Moffitt Cancer Center, n = 59, serum). Logistic regression with backward feature selection on the MD Anderson cohort yielded panels for severe (Grade ≥ 2) CRS and ICANS that were locked and tested without refitting on the Moffitt cohort. Patients were stratified into low-, intermediate-, and high-risk tertiles. Ingenuity Pathway Analysis defined upstream regulators and canonical pathways. The 17 CRS-associated and 21 ICANS-associated consensus proteins were classified into mechanistic themes for biological interpretation.

**Results:** A 5-marker CRS panel (SCRIB, MYL6, MTHFD1L, HSP90B1, MMP2) achieved AUCs of 0.85 (95% CI 0.72–0.98) and 0.76 (0.63–0.89) in the discovery and validation cohorts, respectively. An expanded 8-marker ICANS panel (the CRS panel plus SPOCK2, SLC3A2, CD84) achieved AUCs of 0.91 (0.81–1.00) and 0.67 (0.51–0.84). In the combined dataset, high-risk-tertile patients were 13.84-fold (95% CI 4.21–56.26) and 8.59-fold (2.87–29.09) more likely to develop Grade ≥ 2 CRS and ICANS, respectively. Pathway analysis converged on AKT-driven inflammation and endothelial activation. Functional clustering of the consensus proteins partitioned into mechanistically coherent themes consistent with a dual-anatomy model: severe CRS reflected peripheral macrophage priming and endothelial activation with surplus complement amplification (HSP90B1▴, CSF1▴, MMP2▴, HEG1▴, C3▴) and endotheliopathic coagulation (PROC▾, F7▾), whereas severe ICANS reflected cerebrovascular junction and basement-membrane stripping (CDH5▾, ITGB1▾, FN1▾, brain-enriched SPOCK2▼), hepatic synthetic suppression (TTR▾, APOA2▾, IGFBP3▾), compromised plasma antioxidant capacity (GPX3▾, PON1▾), and inflammasome dis-restraint via DPP9▼. PGLYRP2 and SCRIB depletion were shared by both signatures and identified a common upstream priming substrate.

**Conclusions:** Externally validated, pre-infusion proteomic panels predict severe CRS and ICANS following CAR T-cell therapy and define a coherent pre-infusion endothelial–immune priming axis (HSP90B1, MMP2, AKT) with mechanistically interpretable, druggable nodes. The dual-anatomy framework distinguishes peripheral CRS-biased from cerebrovascular ICANS-biased phenotypes downstream of a shared microbiome-host barrier priming substrate, providing a foundation for biomarker-guided risk stratification and cluster-matched prophylactic intervention to enhance the safety and outpatient feasibility of CAR T-cell therapy.

## Background

Chimeric antigen receptor (CAR) T-cell therapy has transformed the management of refractory lymphoid malignancies [1–6], with seven FDA-approved products now available for hematologic indications. Widespread adoption nevertheless remains constrained by immune-related adverse events, particularly cytokine release syndrome (CRS) and immune effector cell-associated neurotoxicity syndrome (ICANS) [7], whose incidence and severity vary across cellular immunotherapy platforms and represent meaningful threats to patient safety.

Existing predictive models — including machine learning algorithms [8], the EASIX score [9], and post-infusion inflammatory cytokines [10,11] — lack sufficient external validation for routine clinical use, driving universal hospitalization of high-risk patients, escalating costs, and unnecessary empiric antibiotic use [12–14]. As CAR T-cell therapy expands into outpatient settings, rapid, non-invasive, and clinically actionable biomarkers capable of early prediction of CRS and ICANS — both reversible with timely intervention [15] — are urgently needed to enable individualized risk stratification.

Mechanistic studies have implicated host monocyte- and macrophage-derived IL-1 and IL-6, endothelial activation, and disruption of the blood–brain barrier as central effectors of severe CRS and ICANS. Whether these downstream pathways have a measurable pre-infusion biochemical correlate that could enable individualized prophylaxis remains unknown. To address this gap, we applied unbiased mass-spectrometry-based proteomics to biofluids collected immediately prior to CAR T-cell infusion in 98 prospectively-followed patients with relapsed/refractory lymphoma from two academic centers, derived externally validated protein signatures predictive of severe (Grade ≥ 2) CRS and ICANS, defined the underlying mechanistic axes through pathway and theme-based analysis, and integrated the CRS and ICANS signatures into a dual-anatomy framework that links pre-infusion priming biology to syndrome-specific vascular failure.

## Methods

### Study cohorts and biospecimens

Plasma and serum specimens were prospectively collected from patients with relapsed/refractory (r/r) lymphoma who subsequently underwent standard-of-care anti-CD19 CAR T-cell therapy at two academic institutions: The University of Texas MD Anderson Cancer Center (MDACC), Houston, TX, and Moffitt Cancer Center, Tampa, FL. The severity of CRS and ICANS was graded and managed according to American Society for Transplantation and Cellular Therapy (ASTCT) guidelines [7]. All procedures complied with institutional guidelines and clinical data were obtained at each site under local IRB / ethics approvals.

### Proteomics workflow and sample preparation

Proteomic analyses were performed under standard operating procedures in a 96-well high-throughput format. For each specimen, 10 µL of plasma or serum was first incubated with Protein A/G magnetic beads (Thermo Scientific, #PI78610) to capture immunoglobulins and Ig-bound antigens, after which the supernatant underwent high-abundance protein depletion using Hu-14 immunodepletion slurry (Thermo Scientific, #PIA36372) to remove albumin, IgG, IgA, transferrin, haptoglobin, fibrinogen, α1-antitrypsin, α1-acid glycoprotein, apolipoprotein A-I, apolipoprotein A-II, complement C3, transthyretin, IgM, and α2-macroglobulin; the resulting flow-through was used to profile the lower-abundance, non-Ig-bound proteome. Proteins were concentrated, reduced with TCEP (Thermo Fisher Scientific, #77720), alkylated with iodoacetamide (Thermo Scientific, #PIA39271), buffer-exchanged into 50 mM ammonium bicarbonate, digested with trypsin (Thermo Scientific, #PI90059), desalted using a Peptide Clean-Up Plate (Thermo Scientific, #A57865), and dried by SpeedVac. Dried peptides were reconstituted in acetonitrile/water/trifluoroacetic acid (2:98:0.1, v/v/v), quantified by tryptophan fluorescence, and 500 ng were loaded onto EvoTips for LC–MS/MS on an EvoSep system coupled to an 8 cm C18 PepSep column (1.5 µm; Bruker) and analyzed on a timsTOF HT mass spectrometer (Bruker) operated in data-independent acquisition (DIA) mode. RAW files were processed with Spectronaut (Biognosys) against the UniProt Human database (in-silico tryptic digestion; variable PTMs per software defaults) with a 1% false discovery rate; run-to-run quality was monitored by spiking iRT standards (Biognosys) into *Escherichia coli* QC samples to track electrospray stability, retention-time drift, sample loading, signal reproducibility, and peptide identifications across batches.

### Statistical analysis

Discriminative performance of individual proteins and multivariable models was quantified by the area under the receiver operating characteristic curve (AUC), with DeLong’s method used to compute p-values for AUC comparisons and 95% confidence intervals (CIs). Proteins meeting prespecified significance thresholds were submitted to Ingenuity Pathway Analysis (IPA, QIAGEN; version 01-21-03) to identify enriched canonical pathways and upstream regulators. For predictive modeling, we fit logistic regression with backward feature selection on the MDACC plasma cohort (Discovery Set); the final model was locked and evaluated without refitting on the independent Moffitt cohort (Testing Set).

To assess independence from clinical factors, we additionally fit multivariable logistic regression models in which prespecified clinical covariates were entered as confounders alongside proteomic features; model coefficients, standard errors, odds ratios, 95% CIs, and p-values are reported where applicable. Analyses were conducted in R, version 4.2.0. Unless otherwise specified, statistical tests were two-sided with p < 0.05 considered significant.

### Theme-based functional classification of consensus proteins

The 17 CRS-associated and 21 ICANS-associated consensus proteins identified by cross-cohort integration were classified into mechanistic themes through expert curation of UniProt-annotated function, KEGG pathway membership, and prior published roles in vascular, immune, and cerebrovascular biology. Theme assignments were used for biological interpretation only and did not modify the predictive panels. Full theme classification with per-protein direction and brief functional annotation is provided in Supplementary Table S11.

## Results

### Cohort characteristics

The MDACC cohort comprised 39 patients with r/r lymphoma, including 31 with r/r large B-cell lymphoma (LBCL) and 8 with transformed follicular lymphoma (TFL), who had plasma collected on the day of CAR T-cell infusion. The majority of patients received axicabtagene ciloleucel (axi-cel; n = 35), with the remainder receiving tisagenlecleucel (tisa-cel; n = 4). The Moffitt cohort consisted of serum samples from 59 patients with r/r lymphoma, comprising 41 with r/r LBCL, 10 with TFL, and 8 with other r/r lymphomas (Table 1). CAR T-cell products administered in this cohort included axi-cel (n = 38, 64.4%), tisa-cel (n = 19, 32.2%), and lisocabtagene maraleucel (n = 2, 3.4%). In the MDACC cohort, Grade ≥ 2 CRS and ICANS occurred in 15/39 patients (38.5%) and 16/39 patients (41.0%), respectively. In the independent Moffitt cohort, Grade ≥ 2 CRS occurred in 20/59 patients (33.9%) and Grade ≥ 2 ICANS in 18/59 patients (30.5%) (Table 1).

### Discovery of pre-infusion plasma proteomic signatures of severe CRS and ICANS

A total of 1,038 proteins were detected and quantified in plasma samples (Supplementary Table S1). Of these, 58 and 70 were significantly predictive (nominal two-sided p < 0.05) of Grade ≥ 2 CRS and ICANS, respectively (Figure 1A–B), with two proteins (SELPLG and FBLN1) shared between both toxicities (Figure 1C). Ingenuity Pathway Analysis of CRS-associated proteins implicated inflammatory cytokine networks centered on IL-6, with predicted activation of AKT signaling and inhibition of TP53 (Supplementary Figure S2A; Supplementary Tables S2–S3), while ICANS-associated proteins similarly converged on IL-6, TNF, and IFN signaling with activated AKT and ERK as upstream regulators (Supplementary Figure S2B; Supplementary Tables S2–S3).

**Figure 1.**
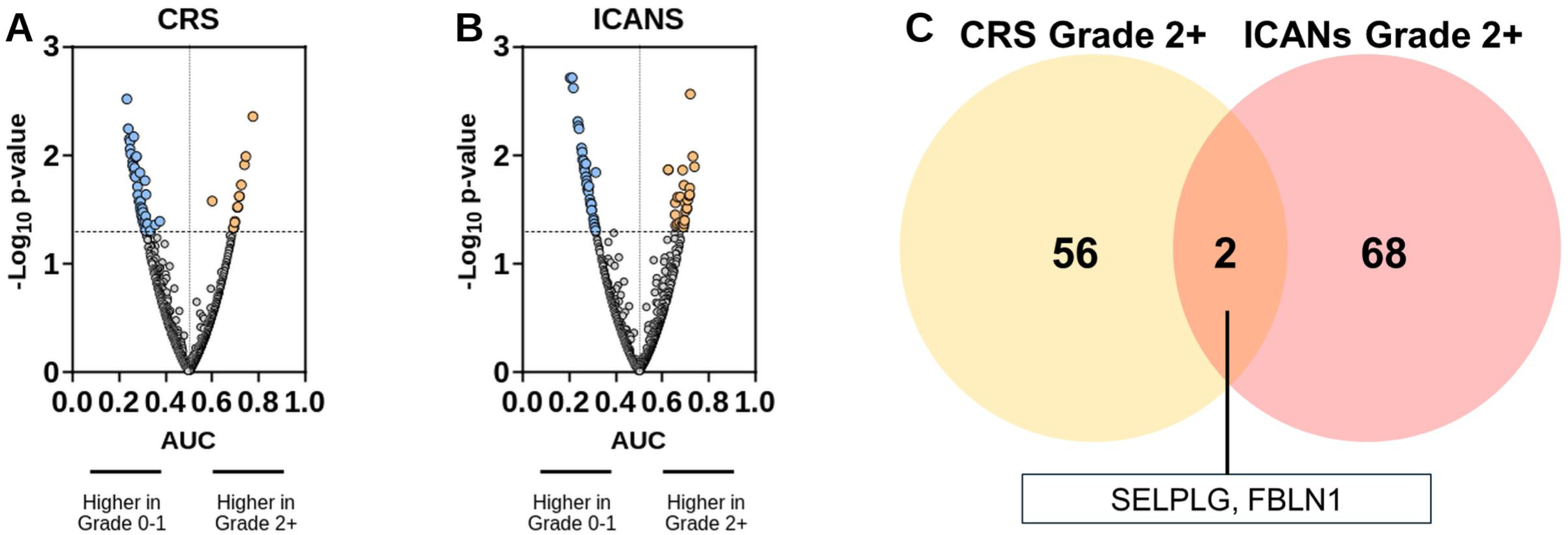
Protein signatures for predicting CAR T-cell–associated CRS and ICANS in the MDACC cohort. (A–B) Volcano plots depicting distribution of quantified proteins for predicting Grade ≥ 2 CRS (A) or Grade ≥ 2 ICANS (B). The x-axis illustrates classifier performance (AUC); the y-axis represents – log of the p-value (two-sided Wilcoxon rank-sum tests). Orange and blue nodes represent protein features that were statistically significantly (nominal two-sided p < 0.05) predictive of CAR T-cell–associated toxicity. (C) Venn diagram showing overlap of proteins predictive of Grade ≥ 2 CRS or ICANS. Networks generated by Ingenuity Pathway Analysis of the CRS- and ICANS-associated protein sets are presented in Supplementary Figure S2 (Supplementary Figures S2A and S2B, respectively).

To validate these findings, proteomic profiling was performed on an independent cohort of 59 r/r lymphoma patients from Moffitt Cancer Center using pre-treatment serum samples. Of 911 proteins quantified, 52 and 33 were predictive of Grade ≥ 2 CRS and ICANS, respectively (Figure 2A–B; Supplementary Table S1), with IPA confirming enrichment of cancer, inflammatory disease, and organismal injury pathways among CRS-associated proteins (Supplementary Tables S4–S5).

**Figure 2.**
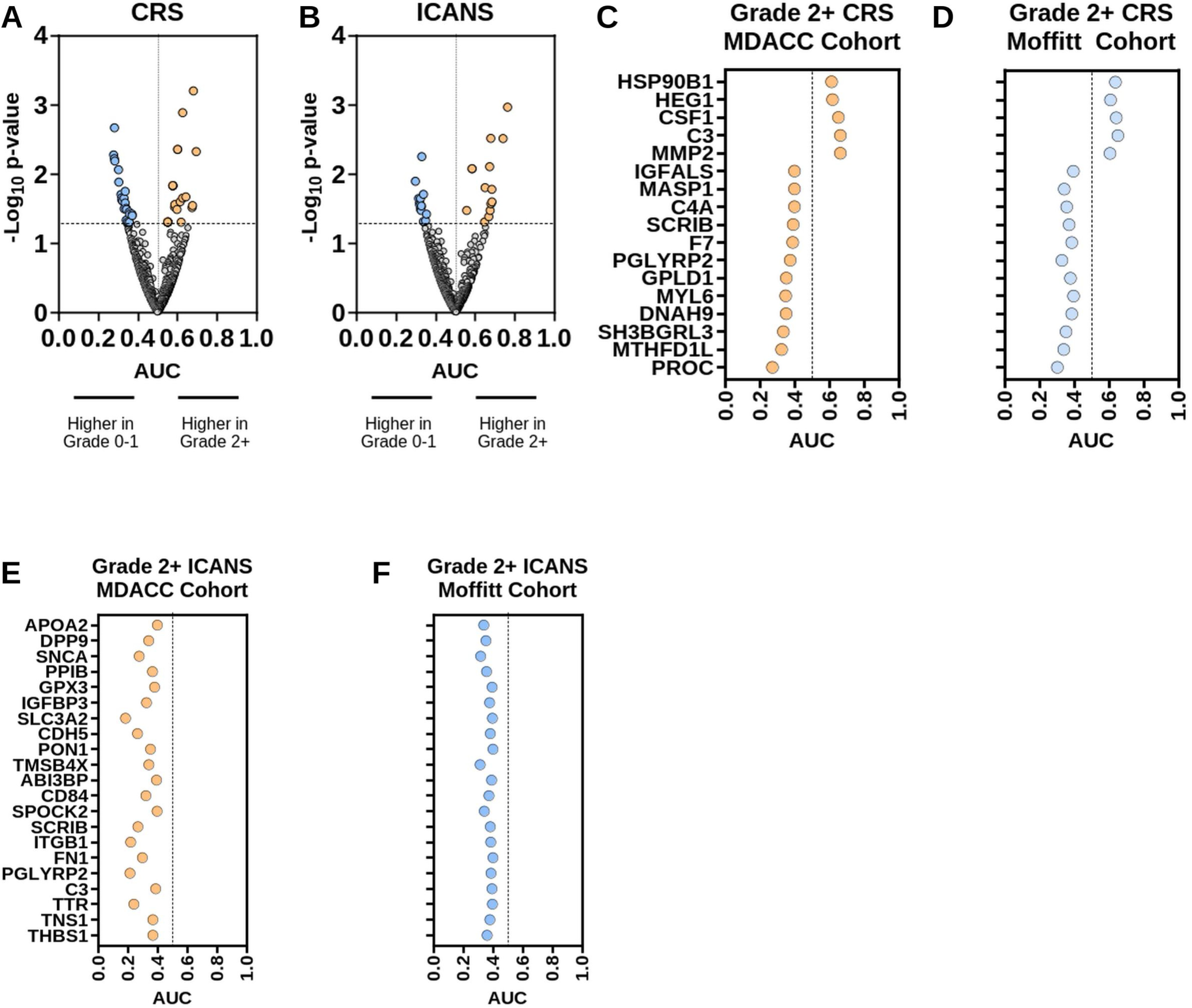
Protein signatures for predicting CAR T-cell–associated CRS and ICANS in the Moffitt cohort. (A–B) Volcano plots depicting distribution of quantified proteins for predicting Grade ≥ 2 CRS (A) or Grade ≥ 2 ICANS (B). (C–D) Dot plots illustrating circulating proteins that were consistently predictive (AUC ≥ 0.60 or ≤ 0.40) of Grade ≥ 2 CRS in the MDACC (C) and Moffitt (D) cohorts. (E–F) Dot plots illustrating circulating proteins that were consistently predictive of Grade ≥ 2 ICANS in the MDACC (E) and Moffitt (F) cohorts. IPA networks generated from the consensus CRS- and ICANS-associated proteins are presented in Supplementary Figure S3.

Cross-cohort integration of the MDACC and Moffitt proteomic profiles identified a consensus signature of 17 CRS-associated proteins, comprising 5 consistently elevated (MMP2, C3, CSF1, HEG1, HSP90B1; AUC ≥ 0.60) and 12 consistently reduced (including PROC, F7, C4A, MASP1; AUC ≤ 0.40, Figure 2C), alongside 21 proteins uniformly downregulated in patients developing Grade ≥ 2 ICANS (Figure 2D). IPA analyses of the 17 CRS-associated and 21 ICANS-associated proteins centered on networks reflecting activation of AKT signaling (Supplementary Figure S3A, S3B).

### Theme-based architecture of CRS- and ICANS-associated consensus proteins

Beyond their convergence on AKT-driven inflammation, the 17 CRS-associated and 21 ICANS-associated proteins partitioned into mechanistically coherent functional clusters that together define two anatomically distinct readouts of a shared upstream priming state. The 17-protein CRS signature segregated into five themes: (i) macrophage priming (CSF1▲, HSP90B1▴, C3▴); (ii) endothelial barrier failure (MMP2▴, HEG1▴, MYL6▾, SCRIB▾); (iii) loss of anti-inflammatory tone (PGLYRP2▼, C4A▾, MASP1▾, GPLD1▾, IGFALS▾); (iv) endotheliopathic coagulation (PROC▾, F7▾); and (v) one-carbon and other metabolic regulators (MTHFD1L▼, SH3BGRL3▾, DNAH9▾). The 21-protein ICANS signature, in contrast, was dominated by protein loss and segregated into seven themes: (i) vascular junction and basement-membrane stripping (CDH5▾, ITGB1▾, THBS1▾, FN1▾, TNS1▾, TMSB4X▼, ABI3BP▼, and the brain-enriched extracellular matrix proteoglycan SPOCK2▼); (ii) hepatic synthetic suppression with a negative acute-phase reactant pattern (TTR▾, APOA2▾, IGFBP3▾, FN1▾, PON1▾); (iii) compromised plasma antioxidant capacity (GPX3▾, PON1▾, APOA2▾); (iv) inflammasome dis-restraint (DPP9▼, a tonic NLRP1 inhibitor); (v) altered blood–brain barrier amino-acid transport (SLC3A2▼, the heavy chain of the LAT1 large-neutral amino-acid transporter); (vi) loss of regulatory tone shared with the CRS signature (PGLYRP2▾, SCRIB▾, CD84▾, PPIB▾); and (vii) a neuronal/synaptic readout (SNCA▼). PGLYRP2▾ and SCRIB▾ were present in both signatures and identify a shared upstream priming substrate, whereas the divergent direction of plasma C3 (▲ in severe CRS, ▼ in severe ICANS) is consistent with site-specific complement engagement: surplus systemic alternative-pathway amplification in severe CRS versus consumptive complement activation at the cerebrovascular endothelium in severe ICANS. Full theme mapping of all 38 proteins is provided in Supplementary Table S11; the dual-anatomy framework is illustrated in Figure 4.

### Development and validation of a 5-marker protein panel for predicting severe CRS

A 5-marker panel for predicting Grade ≥ 2 CRS was developed by applying logistic regression with backward feature selection to the 17 consensus protein biomarkers identified across the MDACC and Moffitt cohorts (Figure 2C). The resulting model, comprising SCRIB, MYL6, MTHFD1L, HSP90B1, and MMP2, achieved an AUC of 0.85 (95% CI 0.72–0.98) for predicting Grade ≥ 2 CRS in the MDACC cohort (Figure 3A).

**Figure 3.**
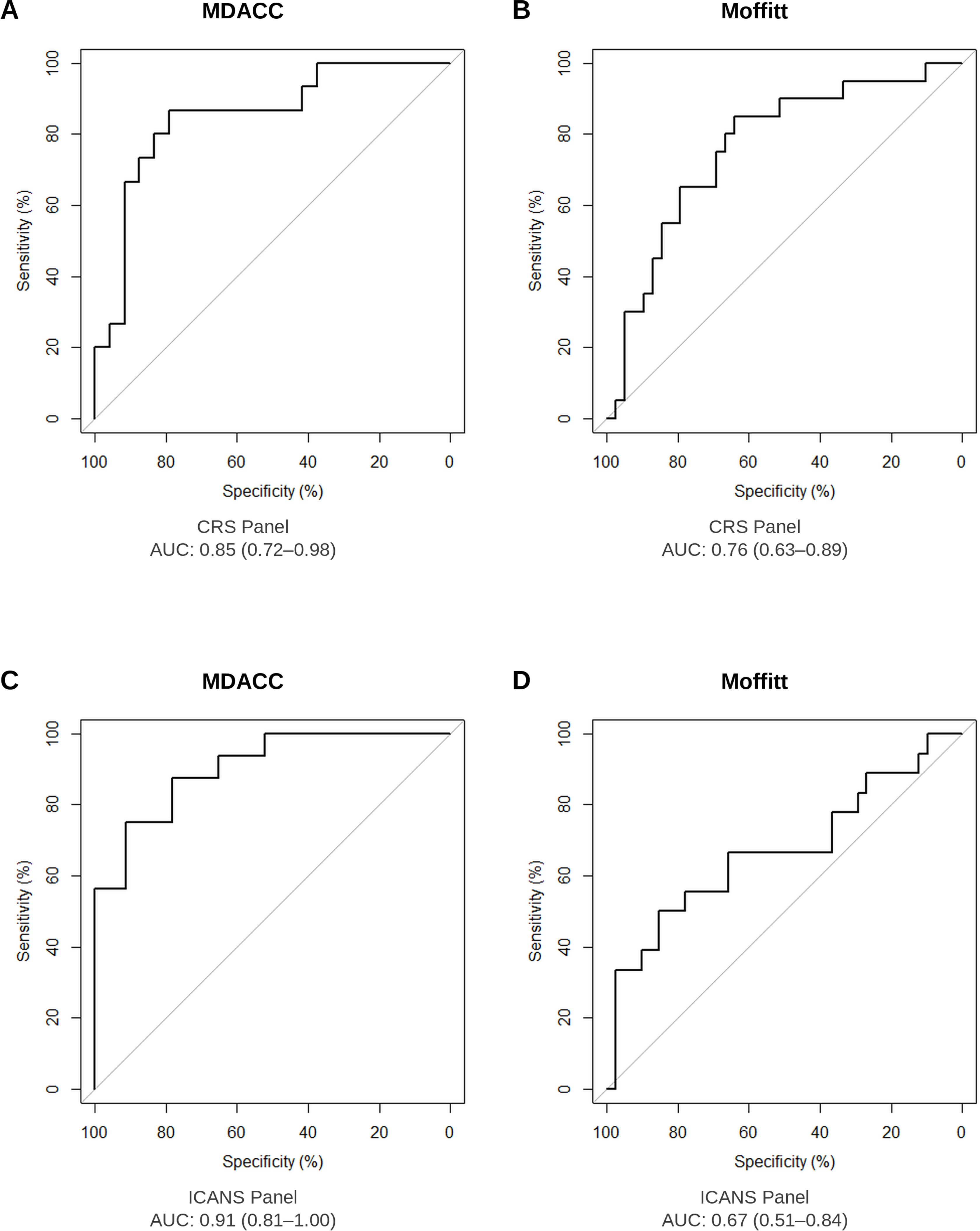
Predictive performance of the CRS panel and ICANS panel for predicting severe (Grade ≥ 2) toxicity following CAR T-cell therapy. (A–B) Classifier performance (AUC) of the 5-marker CRS panel for predicting Grade ≥ 2 CRS in the MDACC (A) and Moffitt (B) cohorts. (C–D) Classifier performance (AUC) of the 8-marker ICANS panel for predicting Grade ≥ 2 ICANS in the MDACC (C) and Moffitt (D) cohorts.

**Figure 4.**
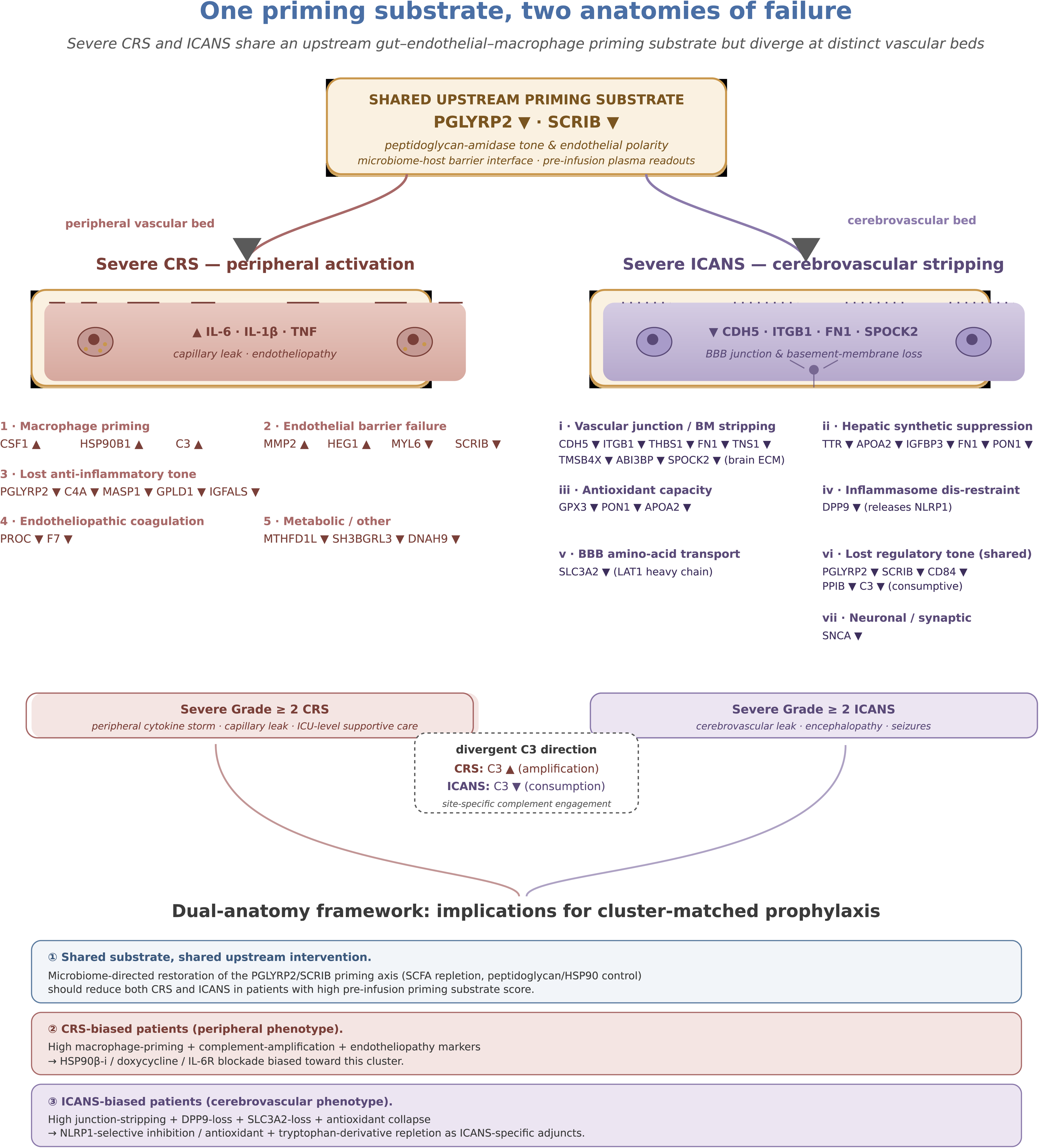
Dual-anatomy framework: one priming substrate, two anatomies of failure. Schematic summarizing the proposed mechanistic model derived from cross-cohort proteomic signatures. A shared upstream pre-infusion priming substrate (PGLYRP2▾, SCRIB▾) bifurcates into two anatomically distinct vascular failure modes. Severe CRS reflects peripheral macrophage activation and endothelial barrier failure with surplus complement amplification (C3▲), endotheliopathic coagulation (PROC▾, F7▾), and macrophage-priming chaperones (HSP90B1▴, CSF1▴). Severe ICANS reflects cerebrovascular junction and basement-membrane stripping (CDH5▾, ITGB1▾, FN1▾, brain-enriched SPOCK2▼) accompanied by hepatic synthetic suppression, compromised plasma antioxidant capacity (GPX3▾, PON1▾), altered blood–brain barrier amino-acid transport (SLC3A2▼), and inflammasome dis-restraint via DPP9▼. The opposite directions of plasma C3 in the two syndromes reflect site-specific complement engagement. The framework supports cluster-matched prophylaxis: shared microbiome-directed restoration of the priming substrate for all high-risk patients, with cluster-specific add-on interventions tuned to whether the patient’s plasma proteome is biased toward peripheral or cerebrovascular failure.

Patients were stratified into low-, intermediate-, and high-risk groups based on panel score tertiles. In the MDACC cohort, patients in the highest-risk tertile (T3) were 18.33-fold more likely to develop Grade ≥ 2 CRS compared to the low-risk reference group (T1; Supplementary Table S6). Multivariable analyses confirmed that the 5-marker panel remained an independent predictor of Grade ≥ 2 CRS after adjustment for sex, age, ECOG performance status, lymphoma subtype, International Prognostic Index (IPI) score, and number of prior therapies (Supplementary Table S7). Subgroup analysis restricted to patients receiving axi-cel (n = 35/39) yielded consistent results (Supplementary Table S8). Notably, in a subset of 24 patients with available cytokine data, cytokine profiles were not significantly associated with Grade ≥ 2 CRS, whereas the 5-marker panel retained its predictive significance (Supplementary Table S9).

External validation of the panel using fixed model coefficients and predefined tertile-based risk strata demonstrated an AUC of 0.76 (95% CI 0.63–0.89) in the independent Moffitt cohort (Figure 3B; Supplementary Table S10). In the combined MDACC and Moffitt dataset, patients in the intermediate-and high-risk strata were 3.15-fold (95% CI 0.92–12.71) and 13.84-fold (95% CI 4.21–56.26) more likely to develop Grade ≥ 2 CRS, respectively, compared to the low-risk group (Table 2).

### Development and validation of an expanded protein panel for predicting severe ICANS

In the MDACC cohort, 16 patients developed Grade ≥ 2 ICANS following CAR T-cell therapy, of whom 10 (62.5%) concurrently experienced Grade ≥ 2 CRS. The 5-marker CRS panel was first evaluated for its capacity to predict Grade ≥ 2 ICANS, yielding an AUC of 0.75 (95% CI 0.59–0.90) (Supplementary Figure S3). To improve predictive sensitivity for ICANS while maintaining clinical feasibility and cost-effectiveness for real-world implementation, we sought to expand the 5-marker CRS panel through the incorporation of additional protein features. Using the continuous 5-marker CRS panel score as a base and applying a backward stepwise selection approach, three additional markers — the proteoglycan SPOCK2, SLC3A2, and CD84 — were incorporated to generate an 8-marker ICANS panel. This expanded panel achieved an AUC of 0.91 (95% CI 0.81–1.00) for predicting Grade ≥ 2 ICANS in the MDACC cohort (Figure 3C). Risk stratification based on panel score tertiles demonstrated that patients in the high-risk stratum were approximately 66-fold more likely to develop Grade ≥ 2 ICANS compared to the low-risk reference group (Supplementary Table S6).

External validation of the 8-marker ICANS panel in the independent Moffitt cohort yielded an AUC of 0.67 (95% CI 0.51–0.84) (Figure 3D). In the combined MDACC and Moffitt dataset, patients in the intermediate- and high-risk strata were 1.21-fold (95% CI 0.36–4.23) and 8.59-fold (95% CI 2.87–29.09) more likely to develop Grade ≥ 2 ICANS, respectively, compared to the low-risk group (Table 2; Supplementary Table S10).

## Discussion

Our study demonstrates that a group of externally validated circulating protein biomarkers measured prior to CAR T-cell therapy can predict the development of severe CRS and ICANS across two independent institutions. Notably, these pre-treatment protein signatures outperformed post-infusion cytokine profiles in a subset of patients, reinforcing the concept that protein-level perturbations precede overt cytokine release and can serve as an earlier, more actionable window for risk stratification. A protein biomarker-driven risk stratification approach could improve the safety of CAR T-cell therapy, reduce ICU admissions, and support its expansion into outpatient settings. Stratifying patients into low-, intermediate-, and high-risk groups could inform decisions regarding prophylactic tocilizumab or corticosteroids, enhanced monitoring intensity, and antibiotic stewardship — the latter being particularly relevant given that antibiotic use may negatively impact gut microbiome composition and potentially modulate immune responses in this vulnerable population. With increased validation of biomarkers, additional targeted drug therapies could be developed, and proteomics technology is anticipated to continue advancing toward real-time testing for use at the clinical bedside [16]. The cross-institutional reproducibility of our panels and the limited number of markers enhance the feasibility of rapid assay development and clinical implementation.

Among the five proteins comprising the severe CRS prediction panel, HSP90B1 (Heat Shock Protein 90 Beta 1), MMP2 (Matrix Metalloproteinase 2), and MYL6 (Myosin Light Chain 6) may have direct mechanistic roles in the pathogenesis of CRS and ICANS. HSP90B1, an endoplasmic reticulum-resident HSP90 isoform, facilitates the proper folding of several immune receptors and signaling molecules, including Toll-like receptors and integrins. HSP90 proteins are highly expressed in lymphoma cells and are known to modulate immune responses by promoting CD8+ T-cell activity [17]. Pharmacological inhibition of HSP90 impairs T-cell proliferation and IFN-γ production [18]. Additionally, HSP90 mediates cytokine production in mast cells via post-transcriptional regulation [19]. Although direct evidence in CRS is limited, it is plausible that HSP90B1 amplifies cytokine storm by sustaining T-cell effector functions, stabilizing key signaling proteins and cytokine mRNAs, and supporting metabolic stress adaptation.

Components of endothelial and extracellular matrix biology are also implicated in the pathogenesis of CRS. MMP2, a matrix metalloproteinase, mediates extracellular matrix remodeling and endothelial activation — both central features of systemic inflammation and vascular permeability [20]. Beyond its role in extracellular matrix remodeling, MMP2 functions as a critical regulator of inflammation-mediated vascular dysfunction, making its pre-treatment elevation a plausible biomarker of a pre-activated endothelial state that predisposes patients to severe CRS [21]. Similarly, MYL6 — a key component of the cytoskeletal actin–myosin complex expressed in vascular smooth muscle and endothelial cells — promotes endothelial activation [22] and aligns with endothelial activation biology, a hallmark of CRS pathology.

Conversely, reduced levels of two proteins — PGLYRP2 (Peptidoglycan Recognition Protein 2) and SCRIB — were consistently associated with severe CRS and ICANS. PGLYRP2 plays a dual role in innate immunity: it hydrolyzes peptidoglycan on bacterial cell wall and modulates inflammation [23]. Because PGLYRP2 cleaves the muramyl-peptide stem of bacterial peptidoglycan and thereby restrains NOD2-driven inflammasome priming, reduced pre-infusion PGLYRP2 may reflect an unchecked gut-derived peptidoglycan / muramyl dipeptide burden, providing a previously unrecognized mechanistic bridge between gut microbiome dysbiosis — which is now established to influence both CAR T-cell efficacy and severe CRS in r/r lymphoma and myeloma — and the endothelial–immune priming state described here. While PGLYRP2 can activate the NOD2–NFκB signaling pathway in macrophages to induce cytokine release, the lower levels in high-risk patients may reflect impaired microbial sensing or dysregulated immune homeostasis. SCRIB, a cell polarity scaffold protein, has been implicated in endothelial inflammatory signaling [24]. Its downregulation may contribute to loss of endothelial barrier integrity — an important feature of severe CRS and exaggerated inflammatory responses. Together, these findings suggest that proteomic signatures preceding CAR T-cell therapy reflect a latent endothelial and immune activation state that predisposes patients to severe immune-related toxicities.

Two of the panel proteins are particularly attractive as direct therapeutic targets. HSP90B1 (GRP94) is required for NLRP3 inflammasome assembly and IL-1β release in macrophages and stabilizes AKT and other signaling clients implicated in IL-6 trans-signaling; isoform-selective HSP90β inhibitors have been shown to reduce LPS-induced TNF-α, IL-1β, and nitric oxide via NF-κB and ERK suppression, and pimitespib is approved for clinical use. MMP2 contributes to glycocalyx shedding and basement-membrane remodeling that mediate capillary leak and blood–brain barrier disruption, and is inhibited at low oral doses by doxycycline, an FDA-approved agent already shown to reduce circulating MMPs and protect the endothelial glycocalyx in cardiopulmonary bypass and acute coronary syndrome trials. Because both axes converge on the AKT signaling hub highlighted by our IPA networks, biomarker-guided pre-infusion endothelial stabilization with these agents represents a near-term, clinically tractable strategy for prophylactic mitigation of severe CRS and ICANS that should be tested in a dedicated mechanistic and clinical pipeline.

A central concern with any prophylactic strategy targeting the inflammatory pathways underlying CRS and ICANS is the potential for impairment of CAR T-cell expansion and anti-lymphoma efficacy, given that IL-6, IL-1, and AKT signaling also support productive T-cell activation. Several lines of evidence argue that the pre-infusion endothelial–immune priming axis can be selectively targeted while preserving efficacy. First, IL-6 receptor blockade with tocilizumab is now standard CRS supportive care and does not impair CAR T-cell cytotoxicity, expansion, or response. Second, in a Phase II of prophylactic anakinra (Park et al., 2023), Day +28 overall response rates were preserved despite IL-1 receptor blockade, demonstrating that selective targeting of macrophage-derived inflammation can dissociate toxicity reduction from response loss. Third, T-cell-intrinsic NOD2 has been shown to be dispensable for CD8+ T-cell immunity, indicating that the adjuvant effect of NOD2/NLRP3 signaling on anti-tumor responses operates primarily through antigen-presenting cells rather than directly on CAR T-cells. Fourth, the cellular and temporal targets of the proposed strategy — doxycycline-mediated endothelial / glycocalyx protection and pre-infusion (rather than on-treatment) HSP90 inhibition — are anatomically and temporally decoupled from the lymph-node and tumor-microenvironment compartments that drive CAR T-cell expansion. Finally, in published gut-microbiome work, the bacterial taxa associated with CAR T-cell response are largely distinct from those associated with severe CRS, and short-chain fatty acid metabolites of barrier-protective commensals enhance CAR T-cell anti-tumor function while protecting endothelial barriers. Together, these observations suggest that toxicity-driving inflammation in heavily-pretreated patients is, at least in part, chronic, sterile, and mistargeted (driven by translocated bacterial PAMPs from a dysbiotic gut barrier) rather than productive anti-tumor inflammation, providing a coherent biological rationale for selective intervention against the priming axis without compromise of efficacy.

Notably, pathway analysis of protein signatures predictive of severe CRS and ICANS converged on enrichment of inflammatory cytokine networks, including IL-6, TNF, and interferons, with predicted activation of downstream PI3K/AKT and ERK signaling cascades. Preclinical models have demonstrated that CRS severity is mediated not by CAR T cell–derived cytokines directly, but by IL-6, IL-1, and nitric oxide produced by recipient macrophages [25]. The interplay between CAR T cells and tumor cells activates host bystander cells, especially macrophages, eliciting distortion of the cytokine network, with massive cytokines subsequently inducing endothelial cell activation and contributing to the constitutional symptoms of CRS [26,27]. While CRS and ICANS share upstream inflammatory drivers, our data suggest divergence in downstream effector mechanisms — with vascular and endothelial activation predominating in severe CRS and neuroinflammatory pathways in ICANS. Microvascular and macrovascular insults, along with white matter lesions observed on imaging in previous studies, suggest that an underlying vascular pathology contributes to secondary CNS injury in ICANS after CRS and CAR T-cell expansion [28]. The presence of these protein-level signals before infusion supports the potential feasibility of prophylactic interventions, suggesting that upstream protein changes may precede cytokine release and offer a more actionable diagnostic and therapeutic window.

Taken together, our cross-cohort proteomic data support a dual-anatomy model in which a shared upstream priming substrate — measurable in pre-infusion plasma as PGLYRP2▾, SCRIB▾, and the broader endothelial–immune priming signature — is read out by two anatomically distinct vascular beds with different vulnerabilities. In patients destined for severe CRS, the dominant phenotype is macrophage activation and peripheral endothelial barrier failure, with surplus systemic alternative-pathway complement amplification (C3▲), endotheliopathic coagulation (PROC▾, F7▾), and macrophage-priming chaperones (HSP90B1▴, CSF1▴) dominating the plasma signal. In patients destined for severe ICANS, the dominant phenotype is cerebrovascular junction and basement-membrane stripping (CDH5▾, ITGB1▾, FN1▾, brain-enriched SPOCK2▼) accompanied by hepatic synthetic suppression, compromised plasma antioxidant capacity (GPX3▾, PON1▾), and altered blood–brain barrier amino-acid handling (SLC3A2▼). The opposite directions of plasma C3 in CRS and ICANS — surplus production versus consumptive loss — and the specifically ICANS-restricted depletion of the inflammasome regulator DPP9 (whose loss disinhibits NLRP1-driven pyroptosis in cells that express NLRP1 most highly, including neurons and brain-resident myeloid cells) provide internally consistent compartmental and mechanistic biomarkers that can be tested in larger cohorts. This framework reconciles why some patients develop primarily CRS, others primarily ICANS, and others both, while preserving a single upstream substrate that is modifiable before CAR T-cell infusion.

Among the ICANS-restricted proteomic signals, depletion of DPP9 deserves particular attention as a previously unrecognized, mechanistically specific therapeutic lead. DPP9 is a tonic intracellular inhibitor of the NLRP1 inflammasome; its functional loss releases NLRP1-driven pyroptosis in cells that express NLRP1 most highly, including neurons and brain-resident myeloid cells. Pre-infusion plasma DPP9 reduction is therefore plausibly both a biomarker of, and a contributor to, the cerebrovascular and neuroinflammatory phenotype that defines ICANS. NLRP1-selective small-molecule inhibitors are entering preclinical development for inflammatory and neurodegenerative indications and represent a near-term, mechanism-aligned candidate for ICANS-specific prophylaxis that, unlike broad cytokine blockade, would be expected to spare productive CAR T-cell expansion. This hypothesis is testable in vitro using human cerebral endothelium–myeloid co-cultures challenged with patient-derived peptidoglycan- and LPS-rich pre-infusion plasma, and in vivo in CAR T-cell xenograft models with DPP9 modulation.

We acknowledge limitations to our study, including modest sample size. Although our statistical models were robust and signatures were replicated across independent cohorts, broader validation across diverse CAR T-cell products, disease contexts, and clinical settings is warranted to confirm performance and define implementation thresholds. Future work will focus on translating these panels into rapid immunoassays, integrating them into clinical workflows, and evaluating their utility in guiding pre-emptive interventions. Mechanistic studies dissecting the roles of HSP90B1, MMP2, MYL6, PGLYRP2, SCRIB, CDH5, SPOCK2, and DPP9 in endothelial, cerebrovascular, and neuroinflammatory circuits are warranted to elucidate the biological basis of these associations and to test the dual-anatomy model in larger cohorts and in mechanistic models.

## Conclusions

We establish externally validated, pre-infusion proteomic panels that predict severe CRS and ICANS following CAR T-cell therapy and identify a coherent endothelial–immune priming axis (HSP90B1, MMP2, AKT) that nominates druggable nodes for prophylactic intervention. The dual-anatomy framework distinguishes peripheral CRS-biased from cerebrovascular ICANS-biased phenotypes downstream of a shared microbiome-host barrier priming substrate, with PGLYRP2 and SCRIB as the shared upstream readouts and DPP9 as a previously unrecognized, ICANS-specific therapeutic lead. Together, these findings provide a foundation both for real-time, biomarker-driven risk stratification and for mechanistically informed, prophylactic therapeutic strategies that may enhance the safety and broader, outpatient adoption of CAR T-cell therapy.

## Data Availability

Data will be available to readers on request ot the author. Most of the data has already been loaded with the manuscript.

## List of abbreviations

ASTCT: American Society for Transplantation and Cellular Therapy
AUC: area under the receiver operating characteristic curve
axi-cel: axicabtagene ciloleucel
BBB: blood–brain barrier
CAR: chimeric antigen receptor
CDH5: cadherin 5 (VE-cadherin)
CI: confidence interval
CRS: cytokine release syndrome
CSF1: colony-stimulating factor 1
DIA: data-independent acquisition
DPP9: dipeptidyl peptidase 9
EASIX: Endothelial Activation and Stress Index
ECM: extracellular matrix
FDR: false discovery rate
GRP94: glucose-regulated protein 94 (HSP90B1)
HDL: high-density lipoprotein
HSP90B1: heat shock protein 90 beta 1
ICANS: immune effector cell-associated neurotoxicity syndrome
IPA: Ingenuity Pathway Analysis
IPI: International Prognostic Index
ITGB1: integrin beta 1
LBCL: large B-cell lymphoma
LC–MS/MS: liquid chromatography–tandem mass spectrometry
liso-cel: lisocabtagene maraleucel
MDP: muramyl dipeptide
MMP2: matrix metalloproteinase 2
MYL6: myosin light chain 6
NLRP1/3: NLR family pyrin domain containing 1/3
NOD2: nucleotide-binding oligomerization domain 2
OR: odds ratio
PAMP: pathogen-associated molecular pattern
PGLYRP2: peptidoglycan recognition protein 2
PROC: protein C
r/r: relapsed/refractory
SCRIB: scribble planar cell polarity protein
SLC3A2: solute carrier family 3 member 2 (CD98 heavy chain)
SPOCK2: SPARC/osteonectin, cwcv and kazal-like domains proteoglycan 2
TFL: transformed follicular lymphoma
tisa-cel: tisagenlecleucel.

## Declarations

### Ethics approval and consent to participate

All study procedures were performed in accordance with the Declaration of Helsinki and applicable institutional and federal guidelines. Plasma collection and clinical data abstraction at The University of Texas MD Anderson Cancer Center were performed under MD Anderson IRB-approved protocols, and serum collection and clinical data abstraction at Moffitt Cancer Center were performed under Moffitt IRB-approved protocols. Written informed consent was obtained from all participants prior to specimen collection.

### Consent for publication

Not applicable. This article does not contain identifiable individual-level data, images, or videos.

### Availability of data and materials

The mass-spectrometry proteomic data generated during this study are available from the corresponding author upon reasonable request and will be deposited in the ProteomeXchange Consortium (PRIDE partner repository) at the time of publication. Source data underlying the panel scoring and risk-stratification analyses are provided in the Supplementary Information. Code used to fit and apply the predictive panels is available from the corresponding author upon reasonable request.

### Authors’ contributions

N.Y.S., T.M.J., M.D.J., J.W., and S.H. conceived and designed the study. N.Y.S., P.S., R.N., D.C., L.E.F., S.A., S.P.I., M.W., F.L.L., M.D., C.R.F., E.S., S.S.N., J.W., M.D.J., and T.M.J. contributed clinical data and biospecimens. J.F.F., H.K., and S.H. performed mass-spectrometry-based proteomic profiling and quality control. E.I. performed statistical analyses, model development, and external validation. N.Y.S. and E.I. interpreted the data and performed theme-based functional classification of consensus proteins. N.Y.S. drafted the manuscript with input from E.I., T.M.J., M.D.J., J.W., and S.H. All authors critically reviewed and approved the final version of the manuscript.

## Acknowledgements

The authors thank the patients and their families for participating in this study, and the clinical and laboratory teams at The University of Texas MD Anderson Cancer Center and Moffitt Cancer Center for their support of specimen collection, processing, and clinical data abstraction.

---

**Table 1. Patient and tumor characteristics for the MDACC and Moffitt cohorts.**

**Table 2. Odds ratios of the 5-marker CRS panel and the 8-marker ICANS panel for Grade ≥ 2 CRS and ICANS among different risk strata in the combined (MDACC + Moffitt) dataset.**

*(Tables 1 and 2 retained from prior version; layout unchanged.)*

