## Supplemental file for "Pre-infusion plasma proteomics identifies an endothelial–immune priming signature predictive of severe cytokine release syndrome and neurotoxicity following CAR T-cell therapy in relapsed/refractory lymphoma"

**SUPPLEMENTAL FIGURES AND TABLES**

**Supplemental Table S1. Comprehensive proteomic profiles of plasma and serum specimens obtained from CAR T-cell recipients across the MDACC and Moffitt Cancer Center cohorts, including all detected and quantified proteins with associated statistical metrics. (provided as an excel sheet attachment)**

**Supplemental Table S2. Top predicted diseases based on IPA analysis of protein biomarkers predictive of Grade 2+ CRS and ICANs in MDACC cohort.**

| **Grade 2+ CRS MDACC Cohort** | |
| --- | --- |
| Top Predicted Diseases | p-value Range |
| Cancer | 4.99E-02 - 1.68E-06 |
| Hematological Disease | 4.59E-02 - 1.68E-06 |
| Immunological Disease | 4.59E-02 - 1.68E-06 |
| Organismal Injury and Abnormalities | 4.99E-02 - 1.68E-06 |
| **Grade 2+ ICANS MDACC Cohort** | |
| Organismal Injury and Abnormalities | 6.35E-03 – 6.83E-10 |
| Infectious Disease | 5.67E-03 – 1.06E-9 |
| Cancer | 6.21E-03 – 2.54E-9 |

**Supplemental Table S3. IPA-based upstream regulator analysis of protein biomarkers predictive of Grade 2+ CRS and ICANs in the MDACC Cohort. (Provided as an excel sheet attachment)**

**Supplemental Table S4. Top predicted diseases based on IPA analysis of protein biomarkers predictive of Grade 2+ CRS and ICANs in Moffitt cohort.**

| **Grade 2+ CRS Moffitt Cohort** | |
| --- | --- |
| Top Predicted Diseases | p-value Range |
| Neurological Disease | 3.68E-02 – 1.67E-07 |
| Organismal Injury and Abnormalities | 3.69E-02 – 1.67E-07 |
| Cancer | 3.69E-02 – 5.81E-07 |
| Inflammatory Disease | 3.50E-02 – 3.59E-06 |
| **Grade 2+ ICANS Moffitt Cohort** | |
| Organismal Injury and Abnormalities | 5.00E-02 – 1.01E-7 |
| Neurological Disease | 5.00E-02 – 5.52E-7 |
| Cancer | 4.89E-02 – 1.45E-6 |

**Supplemental Table S5. IPA-based upstream regulator analysis of protein biomarkers predictive of Grade 2+ CRS and ICANs in the Moffitt Cohort. (Provided as an excel sheet attachment)**

**Supplemental Table S6. Odds Ratios of the 5-marker CRS panel and the 8-marker ICANS panel for Grade 2+ CRS and ICANS among different risk strata in the MDACC Cohort.**

|  | **MDACC Cohort** | | | | | |
| --- | --- | --- | --- | --- | --- | --- |
|  | **CRS** | | | | | |
| Tertile | No CRS | Grade 1 CRS | Grade 2+ CRS | OR | 95% CI | p-value |
| T1 | 1 | 10 | 2 | Reference | | |
| T2 | 1 | 9 | 3 | 1.65 | 0.23-14.57 | 0.62 |
| T3 | 1 | 2 | 10 | 18.33 | 2.99-175.93 | 0.0041 |
|  | **ICANS** | | | | | |
| Tertile | No ICANS | Grade 1 ICANS | Grade 2+ ICANS | OR | 95% CI | p-value |
| T1 | 11 | 1 | 1 | Reference | | |
| T2 | 7 | 2 | 4 | 5.33 | 0.65-114.31 | 0.16 |
| T3 | 1 | 1 | 11 | 66.00 | 7.47-1,657.46 | 0.0012 |

**Supplemental Table S7. Multivariable models evaluating predictive performance of the 5-marker CRS panel for predicting Grade 2+ CRS in the MDACC Cohort when considering pertinent clinical markers as covariates.**

|  | **MDACC Cohort** | | |
| --- | --- | --- | --- |
| **Variable** | **OR** | **95% CI** | **P-value** |
| Sex |  |  |  |
| Female | Reference | | |
| Male | 1.23 | 0.13-16.25 | 0.86 |
| Age | 0.99 | 0.92-1.06 | 0.69 |
| ECOG | 1.47 | 0.39-7.06 | 0.58 |
| Type |  |  |  |
| DLBCL | Reference | | |
| TFL | 0.05 | 0.00-0.88 | 0.1 |
| IPI | 1.64 | 0.46-2.45 | 0.23 |
| Prior Therapies | 1.03 | 0.37-1.57 | 0.94 |
| 5-marker CRS Panel |  |  |  |
| Low-Risk | Reference | | |
| Intermediate-Risk | 1.53 | 0.15-18.39 | 0.72 |
| High-Risk | 21.09 | 2.43-341.63 | 0.012 |

**Supplemental Table S8. Multivariable models evaluating predictive performance of the 5-marker CRS panel for predicting Grade 2+ CRS among the subset of patients (N= 35) that received Axi-cel therapy in the MDACC Cohort.**

|  | **MDACC Cohort** | | |
| --- | --- | --- | --- |
| **Variable** | **OR** | **95% CI** | **P-value** |
| Sex |  |  |  |
| Female | Reference | | |
| Male | 0.77 | 0.07-10.34 | 0.83 |
| Age | 0.93 | 0.83-1.02 | 0.17 |
| ECOG | 1.73 | 0.44-8.74 | 0.45 |
| Type |  |  |  |
| DLBCL | Reference | | |
| TFL | 0.03 | 0.00-0.65 | 0.08 |
| IPI | 1.75 | 0.77-4.26 | 0.18 |
| Prior Therapies | 0.93 | 0.41-2.20 | 0.87 |
| 5-marker CRS Panel |  |  |  |
| Low-Risk | Reference | | |
| Intermediate-Risk | 5.58 | 0.37-122.22 | 0.23 |
| High-Risk | 28.49 | 2.83-621.61 | 0.011 |

**Supplemental Table S9. Odds Ratios (95% Confidence Intervals) of cytokines for predicting Grade 2+ CRS.**

| Variable | OR^ | 95% CI | p-value |
| --- | --- | --- | --- |
| GMCSF | 0.72 | 0.18-1.74 | 0.53 |
| IFN-Y | 1.32 | 0.55-3.56 | 0.51 |
| IL-10 | 1.17 | 0.46-3.13 | 0.70 |
| IL-13 | 0.59 | 0.12-1.50 | 0.36 |
| IL-17A | 0.34 | 0.02-1.17 | 0.23 |
| IL-2 | 0.9 | 0.30-2.14 | 0.82 |
| IL-4 | 0.71 | 0.16-1.74 | 0.52 |
| IL-5 | 0.69 | 0.15-1.71 | 0.51 |
| IL-6 | 1.29 | 0.54-3.29 | 0.55 |
| TNF-a | 0.82 | 0.23-1.97 | 0.69 |
| 5-marker CRS Panel | 6.83 | 1.70-57.09 | 0.028 |

^ per unit standard deviation increase

**Supplemental Table S10. Odds Ratios of the 5-marker CRS panel and the 8-marker ICANS panel for Grade 2+ CRS and ICANS among different risk strata in the Moffitt Cohort.**

|  | **Moffitt Cohort** | | | | | |
| --- | --- | --- | --- | --- | --- | --- |
|  | **CRS** | | | | | |
| Tertile | No CRS | Grade 1 CRS | Grade 2+ CRS | OR | 95% CI | p-value |
| T1 | 3 | 15 | 2 | Reference | | |
| T2 | 2 | 11 | 7 | 4.85 | 0.98-36.25 | 0.0731 |
| T3 | 0 | 8 | 11 | 12.38 | 2.59-93.14 | 0.0042 |
|  | **ICANS** | | | | | |
| Tertile | No ICANS | Grade 1 ICANS | Grade 2+ ICANS | OR | 95% CI | p-value |
| T1 | 12 | 3 | 5 | Reference | | |
| T2 | 11 | 6 | 3 | 0.53 | 0.10-2.53 | 0.43 |
| T3 | 5 | 4 | 10 | 3.33 | 0.89-13.84 | 0.082 |

**Supplemental Table S11. Mechanistic-theme classification of the 17 CRS- and 21 ICANS-associated consensus proteins.**

*Direction (▲/▼) is the change in pre-infusion plasma in patients who subsequently developed Grade ≥2 toxicity, relative to those with Grade <2. Themes are organized to support the dual-anatomy model described in the main text.*

| **Protein** | **Direction** | **Toxicity** | **Mechanistic theme** | **Brief functional annotation** |
| --- | --- | --- | --- | --- |
| **CSF1** | **▲** | CRS | 1. Macrophage priming | M-CSF; macrophage proliferation/activation cue |
| **HSP90B1** | **▲** | CRS | 1. Macrophage priming | GRP94; ER chaperone for TLRs/integrins; required for NLRP3 assembly |
| **C3** | **▲** | CRS | 1. Macrophage priming | Alternative-pathway complement amplification |
| **MMP2** | **▲** | CRS | 2. Endothelial barrier failure | Glycocalyx and basement-membrane proteolysis |
| **HEG1** | **▲** | CRS | 2. Endothelial barrier failure | Heart-of-Glass receptor; endothelial junction shedding |
| **MYL6** | **▼** | CRS | 2. Endothelial barrier failure | Myosin light chain 6; EC contractile/barrier integrity |
| **SCRIB** | **▼** | CRS | 2. Endothelial barrier failure | Polarity scaffold; EC tight-junction maintenance |
| **PGLYRP2** | **▼** | CRS | 3. Lost anti-inflammatory tone | Peptidoglycan amidase; restrains NOD2 inflammasome priming |
| **C4A** | **▼** | CRS | 3. Lost anti-inflammatory tone | Classical complement; immune complex clearance |
| **MASP1** | **▼** | CRS | 3. Lost anti-inflammatory tone | Lectin-pathway pattern recognition serine protease |
| **GPLD1** | **▼** | CRS | 3. Lost anti-inflammatory tone | GPI-PLD; cleaves GPI anchors; modulates inflammation |
| **IGFALS** | **▼** | CRS | 3. Lost anti-inflammatory tone | IGF acid-labile subunit; tissue repair / anabolic tone |
| **PROC** | **▼** | CRS | 4. Endotheliopathic coagulation | Protein C; endothelium-derived anticoagulant; consumed in endotheliopathy |
| **F7** | **▼** | CRS | 4. Endotheliopathic coagulation | Factor VII; consumed in coagulopathic states |
| **MTHFD1L** | **▼** | CRS | 5. Metabolic / other | Mitochondrial one-carbon metabolism |
| **SH3BGRL3** | **▼** | CRS | 5. Metabolic / other | Small adapter; less defined plasma role |
| **DNAH9** | **▼** | CRS | 5. Metabolic / other | Axonemal dynein heavy chain 9 |
| **CDH5** | **▼** | ICANS | i. Vascular junction / BM stripping | VE-cadherin; master endothelial adherens junction |
| **ITGB1** | **▼** | ICANS | i. Vascular junction / BM stripping | β1 integrin; EC–ECM and pericyte adhesion |
| **THBS1** | **▼** | ICANS | i. Vascular junction / BM stripping | Thrombospondin-1; regulates EC quiescence |
| **FN1** | **▼** | ICANS | i. Vascular junction / BM stripping | Fibronectin; basement-membrane / ECM |
| **TNS1** | **▼** | ICANS | i. Vascular junction / BM stripping | Tensin-1; cytoskeletal–ECM linker, focal adhesions |
| **TMSB4X** | **▼** | ICANS | i. Vascular junction / BM stripping | Thymosin β4; actin-sequestering, EC barrier |
| **ABI3BP** | **▼** | ICANS | i. Vascular junction / BM stripping | ABI3 binding protein; ECM |
| **SPOCK2** | **▼** | ICANS | i. Vascular junction / BM stripping | Testican-2; brain-enriched ECM proteoglycan; CSF biomarker |
| **TTR** | **▼** | ICANS | ii. Hepatic synthetic suppression | Transthyretin; classic negative acute-phase reactant |
| **APOA2** | **▼** | ICANS | ii. Hepatic synthetic suppression | Apolipoprotein A-II; HDL component |
| **IGFBP3** | **▼** | ICANS | ii. Hepatic synthetic suppression | IGF binding protein 3; hepatic synthesis |
| **GPX3** | **▼** | ICANS | iii. Plasma antioxidant capacity | Glutathione peroxidase 3; plasma redox buffer |
| **PON1** | **▼** | ICANS | iii. Plasma antioxidant capacity | Paraoxonase-1; HDL-associated antioxidant |
| **DPP9** | **▼** | ICANS | iv. Inflammasome dis-restraint | Dipeptidyl peptidase 9; tonic NLRP1 inhibitor |
| **SLC3A2** | **▼** | ICANS | v. BBB amino-acid transport | CD98 heavy chain (LAT1); BBB neutral amino-acid transport |
| **PGLYRP2** | **▼** | ICANS | vi. Lost regulatory tone (shared) | Peptidoglycan amidase (also in CRS signature) |
| **SCRIB** | **▼** | ICANS | vi. Lost regulatory tone (shared) | Polarity scaffold (also in CRS signature) |
| **CD84** | **▼** | ICANS | vi. Lost regulatory tone (shared) | SLAM-family receptor; myeloid–T crosstalk |
| **PPIB** | **▼** | ICANS | vi. Lost regulatory tone (shared) | Cyclophilin B; ER chaperone |
| **C3** | **▼** | ICANS | vi. Lost regulatory tone (shared) | Complement C3 — note: opposite direction from CRS (consumptive) |
| **SNCA** | **▼** | ICANS | vii. Neuronal / synaptic readout | α-synuclein; predominantly neuronal protein |

**Footnote.**

Six proteins appear in both signatures (PGLYRP2, SCRIB, FN1, APOA2, PON1, C3 — highlighted in ochre under the ICANS section). PGLYRP2 ▼ and SCRIB ▼ define the shared upstream priming substrate. C3 changes in opposite directions in the two syndromes (▲ in severe CRS, ▼ in severe ICANS), consistent with site-specific complement engagement: surplus systemic alternative-pathway amplification in severe CRS versus consumptive complement activation at the cerebrovascular endothelium in severe ICANS.

**Supplemental Figure S1. Top Ingenuity Pathway Analysis (IPA) networks generated from plasma protein biomarkers predictive of Grade 2+ CRS (A) and Grade 2+ ICANS (B) in the MDACC cohort. Node colors represent measured fold-change (red—increased; green—decreased) and predicted activation state (orange—activated; blue—inhibited).**

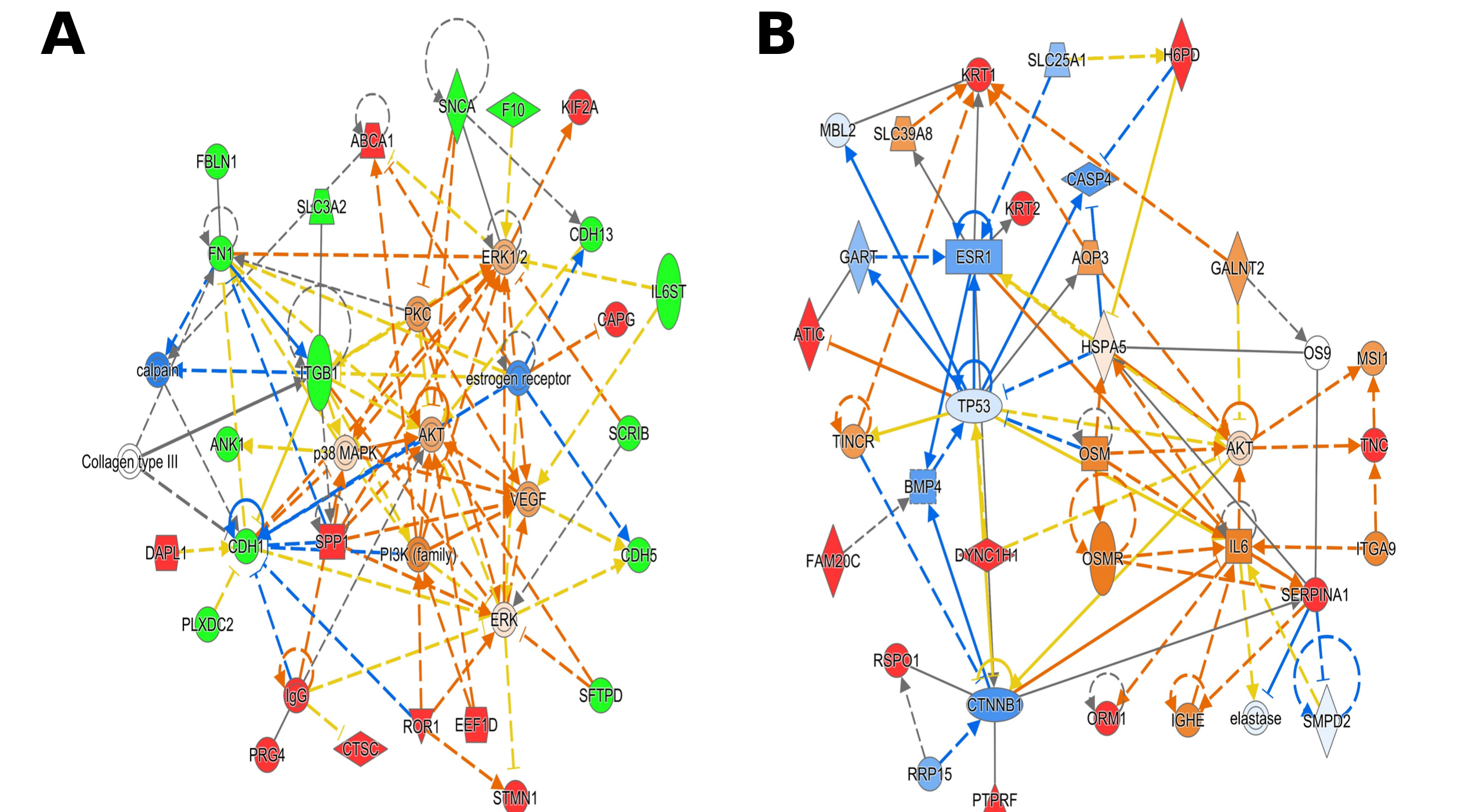

**Supplemental Figure S2. Top Ingenuity Pathway Analysis (IPA) networks generated from plasma protein biomarkers predictive of Grade 2+ CRS (A) and Grade 2+ ICANS (B) in the Moffitt cohort. Node colors represent measured fold-change (red—increased; green—decreased) and predicted activation state (orange—activated; blue—inhibited).**

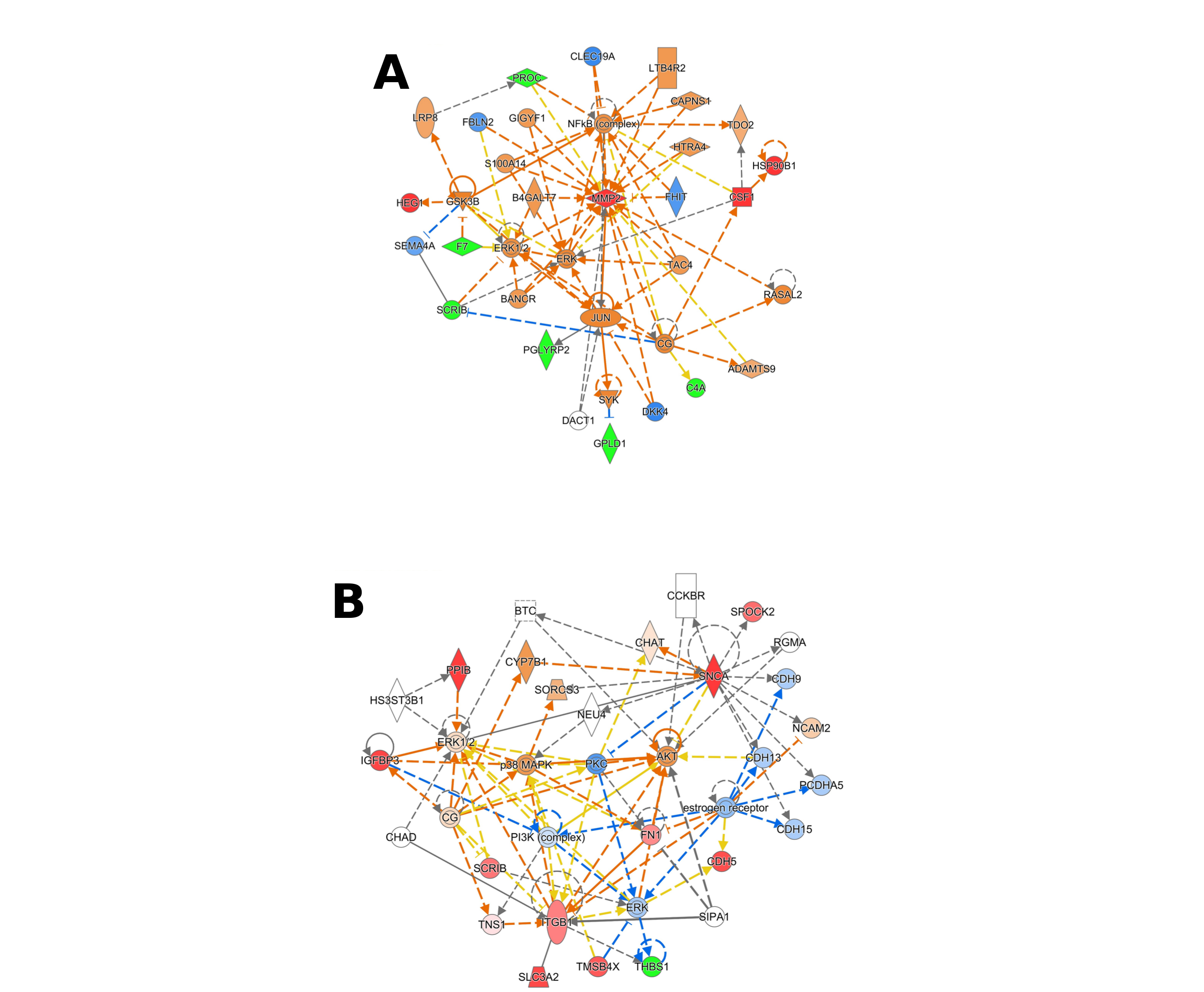

Legend for IPA network nodes and edges (applies to Supplemental Figures S2 and S3).

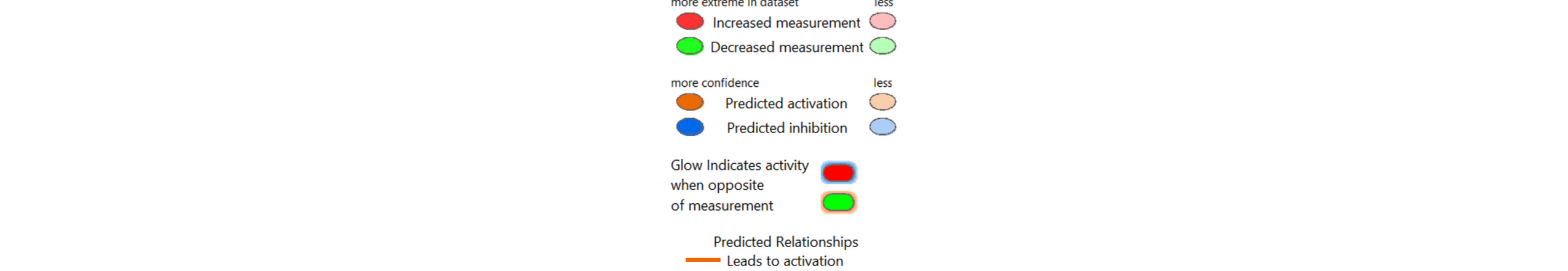

**Supplemental Figure S3. Predictive performance of the 5-marker CRS panel for predicting Grade 2+ ICANS in the MDACC (A) and Moffitt Cohorts (B).**

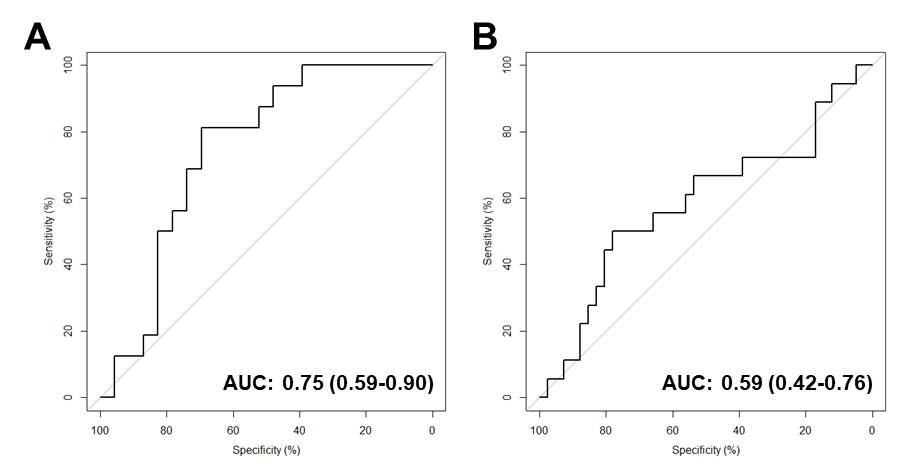
